# Periodontitis and its association with left ventricular geometry and function

**DOI:** 10.1101/2025.07.23.25331894

**Authors:** Maja Trost, Till Ittermann, Birte Holtfreter, Christiane Pink, Marcus Dörr, Henry Völzke, Matthias Nauck, Marcello Ricardo Paulista Markus, Thomas Kocher

**Author notes:** Corresponding author: Prof. Dr. Thomas Kocher, University Medicine Greifswald, Department of Restorative Dentistry, Periodontology, and Endodontology, Fleischmannstr. 42, 17475 Greifswald Germany. E-Mails: Maja Trost, Till Ittermann, Birte Holtfreter, Christiane Pink, Marcus Dörr, Henry Völzke, Matthias Nauck, Marcello Ricardo Paulista Markus, Thomas Kocher.

## Abstract

**Objectives:** The aim of this study was to investigate the cross-sectional association between periodontal status and cardiac structural and functional status, with a focus on left ventricular (LV) geometry and function. The primary research question addressed whether indicators of periodontal disease are linked to markers of LV remodelling.

**Materials and Methods:** Data were drawn from two population-based cohorts in the Study of Health in Pomerania (SHIP): SHIP-START-2 (n=2333) and SHIP-TREND-0 (n=4420). Periodontal status was assessed by bleeding on probing (BOP), probing depth (PD), clinical attachment loss (CAL), number of missing teeth, and the 2018 EFP/AAP classification. LV geometry (end-diastolic/systolic volume, mass, wall thickness) and function (ejection fraction, stroke volume, E/E’ ratio, left atrium diameter) were evaluated using echocardiography in confounder-adjusted regression analyses.

**Results:** Mean PD, CAL, and number of missing teeth were positively associated with end-diastolic volume (PD: β=0.44; 95%CI: 0.13–0.74; CAL: β=0.21; 95%CI: 0.07–0.36; teeth: β=0.06; 95%CI: 0.03–0.09), end-systolic volume (PD: β=0.21; 95%CI: 0.05– 0.37; CAL: β=0.12; 95%CI: 0.05–0.20; teeth: β=0.03; 95%CI: 0.02–0.05), and LV mass (PD: β=1.31; 95%CI: 0.58–2.03; CAL: β=0.52; 95%CI: 0.18–0.86; teeth: β=0.17; 95%CI: 0.10–0.24). Values increased across periodontitis stages (p for trend <0.05). Stage IV was associated with reduced ejection fraction (β=−1.28; 95%CI: −2.45 to −0.12). Other functional markers showed no significant associations.

**Conclusion:** Periodontitis is linked to cardiac remodelling, particularly increased LV volume and mass.

**Clinical Relevance:** Findings support integrating periodontal health into cardiovascular risk management and highlight the need for further research.

## Introduction

According to the World Health Organization, ischemic heart disease and stroke accounted for approximately 13% and 10% of all deaths in 2021, making them the leading causes of death worldwide [1]. Structural changes in the heart are early precursors of coronary heart disease. Left ventricular (LV) volume is a key parameter and independent predictor of coronary morbidity and mortality [2], as it reflects reduced wall elasticity and impaired diastolic function [3]. These changes are major determinants of ejection fraction (EF) and result from hypertension, diabetes, obesity, hyperlipidaemia, or aging [4]. These comorbidities induce a systemic pro-inflammatory state that leads to coronary endothelial inflammation and subsequently contributes to hypertrophy and high diastolic LV stiffness [5]. Left ventricular structural changes are both a consequence and driver of cardiovascular disease, bridging risk factors (e.g. hypertension) and endpoints like heart failure or cardiac death. Structural dimensions are typically assessed by echocardiography (echo).

Periodontitis is a chronic inflammatory disease caused by bacteria that attack and damage the tissues supporting the teeth [6]. In 2017, 9.8% of the world’s adult population, or 796 million people, suffered from severe periodontitis, making it the sixth most common human disease [7]. Apart from dental caries, periodontitis is one of the leading causes of tooth loss [8]. Periodontal bacteria induce local inflammation, with pro-inflammatory mediators and bacteria entering the bloodstream [9]. Thus, periodontitis continuously contributes to low-grade systemic inflammation [10, 11]. Using data from the large-scale Study of Health in Pomerania (SHIP), both periodontitis and low-grade systemic inflammation were shown to be independently and interactively associated with an increased risk of all-cause and cardiovascular disease (CVD) mortality [12]. In addition, a meta-analysis of 48 longitudinal cohorts with 5.71 million participants showed that periodontitis and edentulism increased the risk of CVD mortality [13].

Few publications have investigated whether morphologic changes in the heart are associated with periodontitis. In the Hamburg City Health Study (N=6209), clinically assessed moderate and severe periodontitis was significantly associated only with LV end-diastolic volume fraction, but not with other echocardiographic variables [14]. Similar results have been reported in a small cohort of diabetic patients (N=115) [15]. Another small echocardiographic study (N=66 participants) reported a significant difference in epicardial adipose tissue ratio between the periodontitis and control groups [16]. In SHIP-START-0, women with fewer teeth had higher LV mass (LVM) values than those with more teeth [17]. Using cardiac magnetic resonance imaging measures, a recent study linked self-reported history of periodontitis to interstitial myocardial fibrosis only in male participants of the Multiethnic Study of Atherosclerosis, after adjustment for cardiovascular risk factors and history of myocardial infarction (MI) [18].

As periodontitis affects hypertension [19] and metabolic control [20] and these in turn affect LV structure and function, it is of interest to know, whether periodontitis is an independent risk factor for LV. If so, periodontal treatment, in addition to antihypertensive and antimetabolic treatment, may help to maintain healthy cardiac structure and function. In-depth analyses of periodontitis and structural and functional parameters of the cardiovascular system add to our understanding, as these previous studies reported contradictory results.

Using periodontal and echocardiographic data from two independent population-based studies (SHIP-START-2 and SHIP-TREND-0), we investigated whether periodontal parameters were cross-sectionally associated with structural and functional changes in the heart, particularly LV geometry and function.

## Materials and Methods

### Study design

The SHIP project, conducted in the north-eastern German region of West Pomerania, consists of two independent population-based studies, SHIP-START and SHIP-TREND [21]. SHIP-START baseline participants (N=4308; response 68.8%) were recruited from the general population between 1997 and 2001 aged 20 to 79 years. For this analysis, data were retrieved from SHIP-START-2 (11-year follow-up, 2008-2012, N=2333). SHIP-TREND baseline participants were recruited between 2008 and 2012 from the general population aged 20-79 years (N=4420; response 50.1%).

The final study population of N=4365 participants was derived from the pooled SHIP-START-2 and SHIP-TREND-0 cohorts, after excluding individuals with missing dental (N=676), echocardiographic (N=1,491), or confounder data (N=219). A flowchart illustrating the included and excluded participants of SHIP-START-2 and SHIP-TREND-0 is shown in Figure A1 in the online appendix.

The ethics committee of the University Medicine Greifswald approved SHIP-START-2 (BB 39/08) and SHIP-TREND-0 (BB 39/08a). All participants were informed about the study protocol and signed the informed consent and the privacy statement. The recommendations of the Strengthening the Reporting of Observational Studies in Epidemiology (STROBE) guidelines for observational studies were applied for reporting [22]. The completed STROBE checklist is provided as a supplementary file.

### Periodontitis parameters

Probing depth (PD) and clinical attachment loss (CAL) were measured with a manual periodontal probe (SHIP-START-2: PCP 11; SHIP-TREND-0: PCPUNC 15; Hu-Friedy, Chicago, IL, USA) at four sites per tooth (distobuccal, midbuccal, mesiobuccal, and midlingual) using the half-mouth method, except for the third molars (SHIP-START-2: side alternated; SHIP-TREND-0: side randomized). Measurements were mathematically rounded to the nearest millimetre. The distance between the pocket bottom and the free gingival margin corresponded to the PD.

After periodontal probing, bleeding on probing (BOP) was recorded at the identical four sites on the first incisor, canine, and first molar in each probed quadrant. If an index tooth was missing, the nearest distal tooth was assessed. Details on CAL measurement procedures and exceptions are described in the online appendix.

Periodontal variables included mean PD, mean CAL, the percentage of sites with PD ≥4 mm (%PD≥4mm), percentage of sites with BOP, and the number of missing teeth (excluding third molars). In addition, the EFP/AAP 2018 classification scheme [23] with an adaption of the ACES (Application of the 2018 periodontal status Classification to Epidemiological Survey data) framework for completed studies [24] was applied. Accordingly, PDs (differentiation between stage II and stage III) and the number of opposing pairs of natural teeth (differentiation between stage III and stage IV) were considered as complexity factors. Individuals without teeth were considered edentulous. Individuals with periodontal health, gingivitis, or stage I periodontitis were grouped into the “< stage II” category.

Details on examiner calibration are provided in the online appendix.

### Echocardiography

Two-dimensional, M-mode and Doppler echocardiography was performed using the Vivid-I system (GE Medical Systems, Waukesha, USA). The left ventricle was imaged in M-mode at the papillary level. End-diastolic and end-systolic LV volumes were calculated using the Teichholz equations. LVM was measured in three dimensions using the leading-edge convention [17]. The formulas used to calculate LV wall thickness (LVWT), LV stroke volume (SV), LVEF, and left atrium diameter (LAD) were in accordance with the American Society of Echocardiography guidelines [25]. Interobserver agreement was >90% in the certification study [26]. The following measures were normalized to the allometric power of 2.7, which linearizes the relationship between the variable and height and determines the effect of obesity: LV end-diastolic volume, LV end-systolic volume, LV end-diastolic wall thickness, LV end-diastolic mass, LVSV, and LAD indexed to height in meters [27]. Peak diastolic velocity of early (E) mitral inflow was measured by trans-mitral pulsed-wave Doppler. The E to E’ ratio was calculated by combining this measurement with the tissue Doppler mean (derived from the lateral and medial measurements) of the early (E’) mitral annular peak diastolic velocity [25].

### Covariates

Detailed information on covariates can be found in the online appendix.

### Statistical analyses

Characteristics of the study population were stratified by the 2018 classification and presented as medians, 25^th^ percentiles and 75^th^ percentiles for continuous data or as absolute numbers and percentages for categorical data. Associations of periodontal variables with parameters of echocardiography were analysed by linear regression models adjusted for age, sex, smoking status, known or diagnosed type 2 diabetes, years of schooling, number of dental visits, fat mass, fat-free mass, history of MI, and study. Confounders were chosen based on clinical knowledge. In this study, hypertension was classified as a mediator rather than a confounder of periodontitis, because periodontitis can lead to hypertension via inflammatory mechanisms, which then further increases the risk of cardiovascular disease [28, 29]. Results were reported as β coefficients, 95% confidence intervals (95%CI) and p-value. For the 2018 classification stages, we also calculated a p for trend by introducing this exposure as a continuous variable in the linear regression model. Based on prior knowledge, non-classified cases were ranked between stage IV and edentulous cases. We conducted sensitivity analyses in individuals without a history of MI (N=146). A p<0.05 was considered statistically significant. Calculations were done with Stata 18.5 (Stata Corporation, TX, USA).

## Results

### Baseline characteristics

Overall, 17.1% of participants were categorized into “< stage II”, 17.2% had stage IV periodontitis, and 5.4% were edentulous (Table 1). Individuals who scored higher in the 2018 classification scheme tended to be older, more often male, less educated, and had higher BMI, waist circumference, fat, and fat-free mass. Also, they were more likely to have hypertension, diabetes, and MI. The worse the periodontitis stage, the worse the echocardiographic measurements (left end-diastolic volume, LVWT, LVM, SV, EF, and LAD). For example, LVM ranged from 251 g in “< stage II” to 296 g in stage IV cases. For non-classified cases, echocardiographic measurements were mostly between the stage IV and edentulous cases.

**Table 1.**
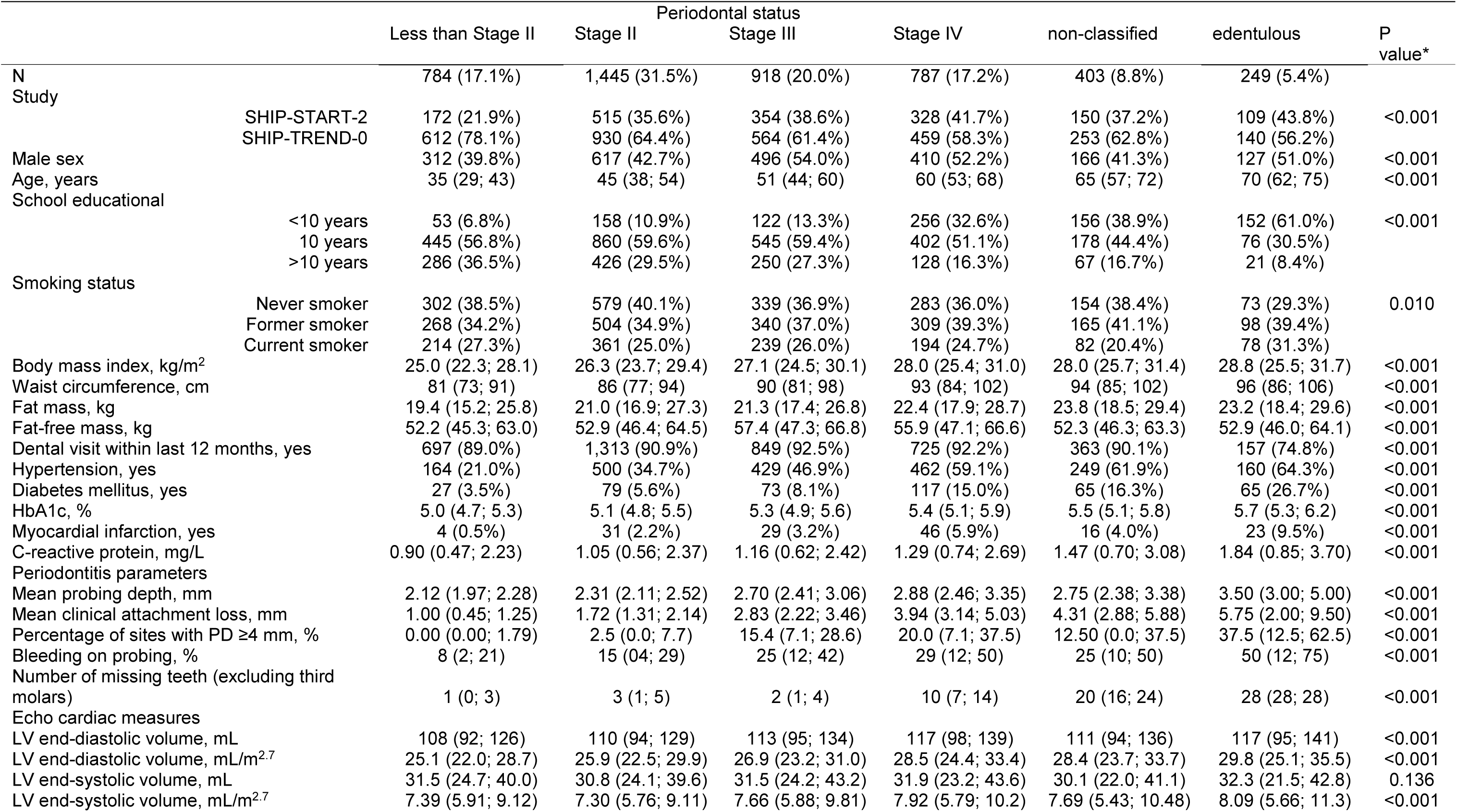
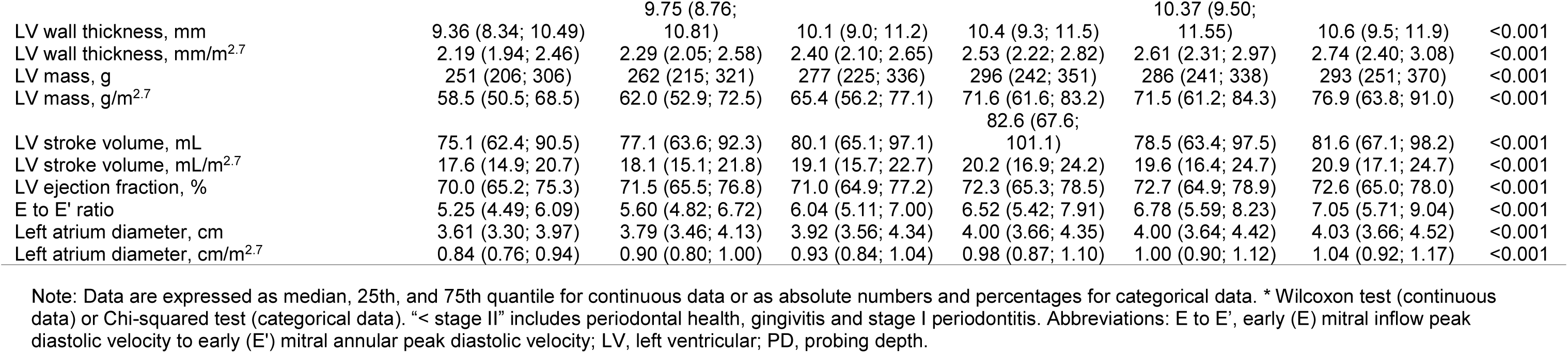
Characteristics of the study population stratified by periodontal status.

### Associations of periodontitis parameters with left ventricular geometry

After adjustment, mean PD and the number of missing teeth were significantly associated with all LV geometry variables (end-diastolic volume, end-systolic volume, LVWT, and LVM). All periodontitis variables except BOP and %PD≥4mm were consistently positively associated with end-diastolic volume (mean PD: β=0.44 (95%CI: 0.13; 0.74), mean CAL: β=0.21 (95%CI: 0.07; 0.36), number of missing teeth: β=0.06 (95%CI: 0.03; 0.09)) and end-systolic volume (mean PD: β=0.21 (95%CI: 0.05; 0. 37), %PD≥4mm: β=0.01 (95%CI: 0.00; 0.01), mean CAL: β=0.12 (95%CI: 0.05; 0.20), number of missing teeth: β=0.03 (95%CI: 0.02; 0.05)) and LVM (mean PD: β=1. 31 (95%CI: 0.58; 2.03), %PD≥4mm: β=0.03 (95%CI: 0.01; 0.06), mean CAL: β=0.52 (95%CI: 0.18; 0.86), the number of missing teeth: β=0.17 (95%CI: 0.10; 0.24)). A 1 mm higher mean PD was associated with a 0.44 (ml/m^2.7^) (95%CI: 0.13; 0.74) higher end-diastolic volume, whereas a 1 mm higher mean CAL was associated with a 0.21 (ml/m^2.7^) (95%CI: 0.07; 0.36) higher end-diastolic volume. A similar association was observed for end-systolic volume (mean PD: β=0.21 (95%CI: 0.05; 0.37), CAL: (β=0.12 (95%CI: 0.05; 0.20)) and LVM (mean PD: β=1.31 (95%CI: 0.58; 2.03), CAL: (β=0.52 (95%CI: 0.18; 0.86)). Each missing tooth contributed to a higher value in end-diastolic volume of 0.06 ml/m^2.7^ (95%CI: 0.03; 0.09).

When evaluating the 2018 classification (Table 3), only stage IV cases and edentulous cases had significantly higher end-diastolic (stage IV: β=1.07 (95%CI: 0.35; 1.80), edentulous β=1.66 (95%CI: 0.56; 2.76)) and end-systolic volumes (stage III: β=0.04 (95%CI: 0.06; 0.74), stage IV: β=0.69 (95%CI: 0.31; 1.08), non-classified: β=0.75 (95%CI: 0.29; 1.21), edentates: β=0.96 (95%CI: 0.38; 1.55)) and a higher LVM (stage IV: β=2.16 (95%CI: 0.45; 3.87), edentates: β=4.04 (95%CI: 1.45; 6.64)) compared to <stage II individuals. No significantly higher results were observed in participants with stage II and III periodontitis compared to reference individuals. End-diastolic volume, end-systolic volume, and LVM showed linearly higher values across categories of the 2018 classification scheme (p for trend <0.05). LVWT was not significantly associated with any of the periodontitis variables. Adjusted means of end-systolic volume index and LVM index for different levels of mean PD, mean CAL, number of missing teeth, and 2018 classification are shown in Figure 1.

**Fig. 1.**
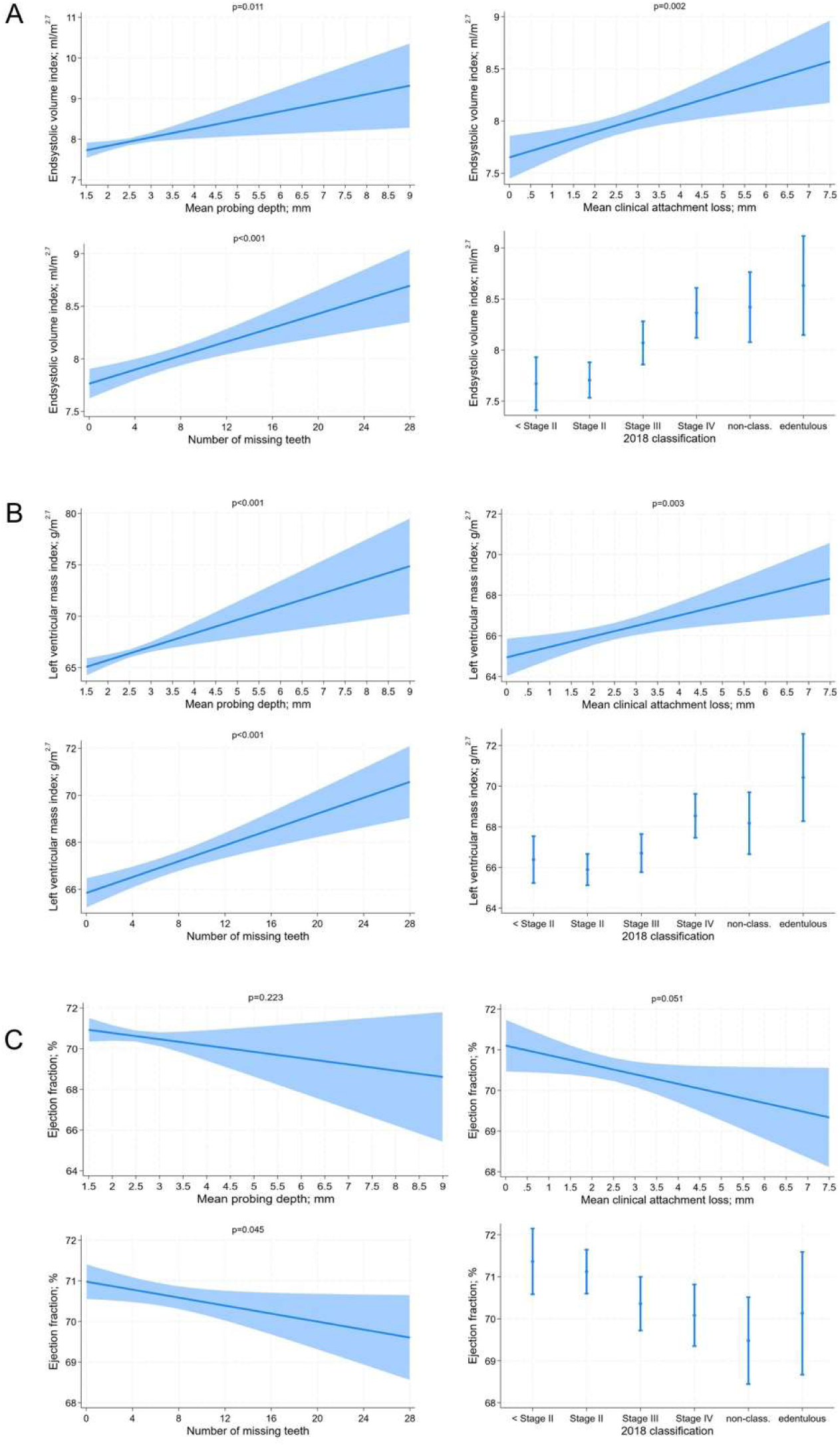
Adjusted means, with 95% confidence intervals, of (**A**) end-systolic volume index (ml/m^2.7^) for mean probing depth (p=0.011), mean clinical attachment loss (p=0.002), the number of missing teeth (p<0.001), and the 2018 classification (p for trend <0.001); (**B**) left ventricular mass index (g/m^2.7^) and mean probing depth (p=0.001), mean clinical attachment loss (p=0.003), the number of missing teeth (p<0.001), and the 2018 classification (p for trend <0.001); (**C**) ejection fraction (%) and mean probing depth (p=0.223), mean clinical attachment loss (p=0.051), the number of missing teeth (p=0.045), and the 2018 classification (p for trend=0.004). “< stage II” includes periodontal health, gingivitis and stage I periodontitis. Models were adjusted for age, sex, school education, smoking status, known or diagnosed diabetes, dental visits within the last 12 months, fat mass, fat-free mass, myocardial infarction and study.

### Associations of periodontitis parameters with left ventricular function

There were weak associations between LVEF and the %PD≥4mm (β=−0.02 (95%CI: −0.04; −0.00)) and the number of missing teeth (β=−0.05 (95%CI: −0.10; −0.00)) (Table 2). SV and LAD were not significantly associated with any of the periodontal variables. BOP was positively associated with the E to E’ ratio.

**Table 2.** Results from linear regression models for associations of periodontitis parameters with cardiac measures (normalized to the height^2.7^) using pooled data from the Studies of Health in Pomerania.

| Outcome variable | BOP |  | Mean PD |  | Percentage of sites with PD ≥ 4mm |  | Mean CAL |  | Number of missing teeth |  |
| --- | --- | --- | --- | --- | --- | --- | --- | --- | --- | --- |
|  | β (95% CI) | P value | β (95% CI) | P value | β (95% CI) | P value | β (95% CI) | P value | β (95% CI) | P value |
| LV end-diastolic volume; mL/m<sup>2.7</sup> | 0.56 (-0.24; 1.36) | 0.169 | 0.44 (0.13; 0.74) | 0.006 | 0.01 (-0.00; 0.02) | 0.099 | 0.21 (0.07; 0.36) | 0.004 | 0.06 (0.03; 0.09) | <0.001 |
| LV end-systolic volume; mL/m<sup>2.7</sup> | 0.38 (-0.04; 0.80) | 0.074 | 0.21 (0.05; 0.37) | 0.011 | 0.01 (0.00; 0.01) | 0.012 | 0.12 (0.05; 0.20) | 0.002 | 0.03 (0.02; 0.05) | <0.001 |
| LV wall thickness; mm/m<sup>2.7</sup> | 0.042 (-0.011; 0.095) | 0.117 | 0.023 (0.002; 0.043) | 0.030 | 0.001 (-0.000; 0.001) | 0.062 | 0.005 (-0.005; 0.014) | 0.348 | 0.002 (0.000; 0.004) | 0.018 |
| LV mass; g/m<sup>2.7</sup> | 1.88 (0.00; 3.75) | 0.050 | 1.31 (0.58; 2.03) | 0.001 | 0.03 (0.01; 0.06) | 0.017 | 0.52 (0.18; 0.86) | 0.003 | 0.17 (0.10; 0.24) | <0.001 |
| LV stroke volume; mL/m<sup>2.7</sup> | 0.18 (-0.50; 0.85) | 0.604 | 0.22 (-0.04; 0.48) | 0.093 | 0.00 (-0.01; 0.01) | 0.697 | 0.09 (-0.03; 0.22) | 0.143 | 0.02 (-0.00; 0.05) | 0.082 |
| LV ejection fraction; % | -0.87 (-2.16; 0.41) | 0.183 | -0.31 (-0.81; 0.19) | 0.223 | -0.02 (-0.04; -0.00) | 0.046 | -0.24 (-0.47; 0.00) | 0.051 | -0.05 (-0.10; -0.00) | 0.045 |
| E to E' ratio | 0.28 (0.08; 0.50) | 0.008 | 0.04 (-0.04; 0.12) | 0.327 | 0.00 (-0.00; 0.00) | 0.524 | -0.01 (-0.05; 0.02) | 0.474 | 0.00 (-0.01; 0.01) | 0.680 |
| Left atrium diameter; cm/m<sup>2.7</sup> | 0.006 (-0.014; 0.025) | 0.567 | 0.002 (-0.005; 0.010) | 0.544 | 0.000 (-0.001; 0.000) | 0.585 | 0.003 (-0.001; 0.006) | 0.139 | 0.000 (-0.001; 0.001) | 0.923 |

Models were adjusted for age, sex, school education, smoking status, known or diagnosed diabetes, dental visits within the last 12 months, fat mass, fat-free mass, myocardial infarction and study. Abbreviations: β, beta regression coefficient; BOP, Bleeding on probing; CAL, clinical attachment loss; CI, confidence interval; E to E', early (E) mitral inflow peak diastolic velocity to early (E') mitral annular peak diastolic velocity; LV, left ventricular; PD, probing depth.

For the 2018 classification (Table 3, Figure 1), EF was significantly lower in stage IV cases (β=−1.28 (95%CI: −2.45; −0.12)) and non-classified individuals (β=−1.89 (95%CI: −3.28; −0.49)); p for trend 0.004. SV, E to E’ ratio, and LAD were not significantly associated with the 2018 classification.

**Table 3.**
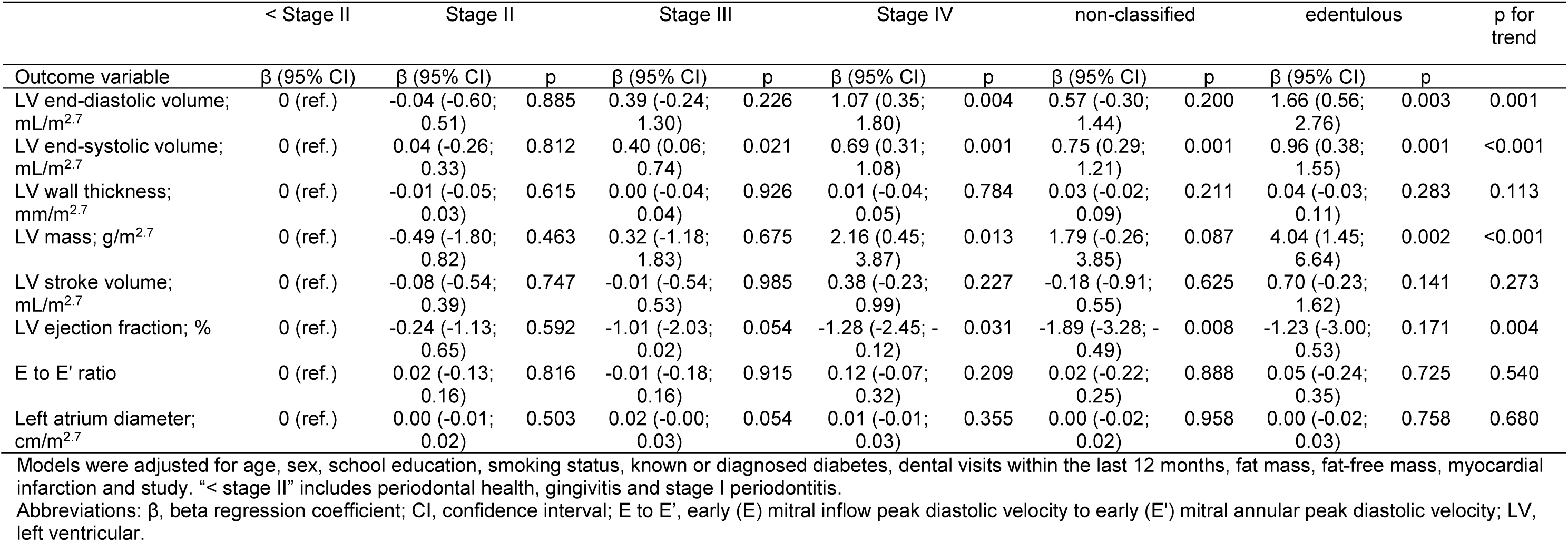
Results from linear regression models for associations of the 2018 classification (ref. “< stage II” (periodontal health, gingivitis or stage I periodontitis)) with cardiac measures (normalized to height^2.7^) using pooled data from the Studies of Health in Pomerania.

Sensitivity analyses were performed excluding participants with a history of MI to assess the reliability of the results for LV geometry and function. The results of the analyses are presented in the online appendix, tables A1 and A2. These analyses showed that the exclusion of MI cases did not significantly alter the observed associations, providing further evidence of the stability of the findings.

## Discussion

This study investigated the relationship between periodontitis and cardiac remodelling. Periodontitis, especially in advanced stages, was significantly correlated with significant changes in cardiac geometry, while its effect on cardiac function appeared to be less pronounced. We observed that participants with severe periodontitis had higher end-systolic volume and LVM, suggesting a compensatory response to the underlying inflammation associated with periodontitis.

Although there is evidence of an association between periodontal disease and CVD, the exact process has not been clearly established [9]. Either periodontopathic infections entered the bloodstream directly, or they indirectly increased the concentration of inflammatory mediators in the bloodstream [30, 31] and persistent low-grade systemic inflammation was exacerbated by periodontitis [12]. Chronic inflammation potentially led to systemic effects, including endothelial dysfunction and increased vascular resistance, which may have contributed to cardiac remodelling [5]. Our data supported this notion, as higher stages of periodontitis were associated with less favourable echocardiographic measures.

Notably, the associations of periodontal parameters with anatomical changes were significant. However, the associations with functional parameters such as LVEF and SV were weaker and less pronounced. This suggested that although the heart underwent structural changes in response to periodontal inflammation, it retained its functional capacity, at least in the early to intermediate stages of periodontitis. One reason is that the structural changes seen are so small that they do not result in functional deficits detectable by the available measurements. For example, a 1 mm change in PD correlates with a 0.44 ml/m^2.7^ change in LV end-diastolic volume, which is a marginal effect with an average LV size of 25-29 ml/m ^2.7^. Similarly, a 1 mm change in PD results in a 1.31 g change in LVM, which is a minimal effect with a baseline mass of 60-70 g. From a statistical perspective, we cannot distinguish between a true lack of physiological effect and an inability to detect subtle changes. Furthermore, the lack of functional damage may indicate that the heart effectively compensates for these minimal structural changes [32]. Other risk factors (e.g. hypertension, diabetes mellitus) can also have a structural effect on the heart muscle without immediately leading to functional impairment [33–35].

In addition, our analysis revealed that the severity of periodontitis was associated with progressively less favourable cardiac remodelling irrespective of considered variable (PD, CAL, number of missing teeth, 2018 classification). The higher the stage of periodontitis was, the higher the effect estimates of echocardiographic parameters, particularly in the transition from healthy or mildly affected to severe periodontitis. As discussed above, the structural effects of periodontitis became significant at higher stages. These findings suggest that although periodontitis affects the cardiac structure, its effects remain subtle and become more pronounced only at advanced stages, which may explain the lack of significant associations with functional changes.

The non-classified cases represent a group that was introduced by the ACES framework [24]. This group includes subjects who could not be classified according to the periodontitis case definition [23], for example, subjects with crowned anterior teeth on which CAL could not be measured, or subjects with only one or two adjacent remaining teeth. We placed unclassified cases between stage IV and edentulous cases (Table 1) because both their oral and general health status were between these two groups, including cases with only a single remaining tooth (with CAL measurements) or two adjacent remaining teeth, allowing CAL and PD measurements. On average, they had more missing teeth (20) than the stage IV group (10). In line, their average HbA1c (5.5%) was between the healthier groups (5.0-5.4%) and the edentulous group (5.7%). The prevalence of hypertension (61.9%) and diabetes (16.3%) was higher than in stage IV, but lower than in the edentate. Based on these observations, we believe that we should not ignore this group and that we have correctly placed them between stage IV and edentulous.

Edentates had significantly higher end-diastolic and end-systolic volumes, suggesting that the cumulative effect of periodontitis and other oral conditions, such as caries, that eventually lead to tooth loss may have further exacerbated cardiac remodelling. While previous studies have shown that the risk of heart disease is associated with the number of missing teeth [36, 37], we did not observe a notable effect on functional remodelling. In our data, LV remodelling was extensive after MI. Although LV remodelling occurred extensively after MI, exclusion of participants with MI did not change our results. Whether this observation is due to reduced performance, a small change in remodelling, or whether the heart was able to compensate for the changes in geometry is an open question.

It is important to mention the strengths and limitations of our study. First, the large sample size. Second, we comprehensively evaluated the associations of various periodontitis parameters with multiple echocardiographic measures, which allowed a nuanced understanding of the association of periodontitis with cardiovascular remodelling. This, together with the use of standardized definitions of periodontitis severity and extent increased the validity of our results.

Limitations include the cross-sectional study design, which limits our ability to draw causal conclusions. Longitudinal studies would have been beneficial to elucidate the temporal relationship between periodontitis and cardiac changes. Second, the results may not be generalizable to other populations. Third, periodontal measurements were recorded using a half-mouth protocol, which is known to underestimate periodontal disease prevalence and severity [38]. However, for measurements such as mean PD/CAL, the level of bias is nevertheless small. Effect estimates are generally shifted towards the null effect [39]. Finally, although we controlled for several potential confounders, residual confounding may still have been an issue, as certain factors that could influence the outcomes were not addressed in this study.

## Conclusion

In conclusion, our findings underscore the importance of considering periodontitis as an independent risk factor for cardiac remodelling. The observed associations highlight the need for further research to understand the underlying mechanisms and to investigate whether interventions targeting periodontal disease may have beneficial effects on cardiovascular health.

## Authors contributions

TK, BH, MRPM substantially contributed to the conception or design of the work. MT, TK, TI, BH, CP, MRPM contributed to the analysis, or interpretation of data. MT, BH, MRPM and TK drafted the work. CP, MD, HV and MN revised the work critically for important intellectual content. All authors approved the final version of the manuscript and are accountable for all aspects of the work.

## Data availability

The data that support the findings of this study are available from Forschungsverbund Community Medicine. Restrictions apply to the availability of these data, which were used under license for this study. Data are available from https://transfer.ship-med.uni-greifswald.de/FAIRequest/login with the permission of Forschungsverbund Community Medicine.

## Ethical approval statement

All SHIP studies were positively evaluated by the ethics committee of the University of Greifswald (SHIP-START-2: BB 39/08; SHIP-TREND-0: BB 39/08a). All participants were informed about the study protocol and signed the informed consent and the privacy statement.

## Online Appendix

### Materials and Methods

#### Periodontitis parameters

##### CAL measurement

If recession was present, CAL was measured directly as the distance between the pocket bottom and the cementoenamel junction (CEJ). If the gingiva covered the CEJ, CAL was measured as the PD minus the distance between the free gingival margin and the CEJ. CAL was not assessed in cases where the determination of the CEJ was unclear (wedge-shaped defects, fillings, and crown margins).

#### Examiner calibration

Dental examinations were performed by five trained and calibrated dentists, with SHIP-START-2 and SHIP-TREND-0 running concurrently. In calibration exercises, all dentists repeatedly examined five individuals not involved in the study. Intra-rater correlations for CAL measurements ranged from 0.67 to 0.89, and the inter-rater correlation was 0.70. For PD measurements, the examiners obtained intra-rater correlations ranging from 0.68 to 0.88 and an inter-rater correlation of 0.72. For the assessment of dental status, Cohen’s kappa reliability coefficients were 0.93-0.99 (intra-examiner) and 0.94-0.98 (pairwise inter-examiner).

#### Covariates

Computer-assisted personal interviews were used to collect data on sex, sociodemographic characteristics (school education; <10/10/>10 years), smoking habits (never smoker, former smoker, current smoker), dental visits within the last 12 months (yes, no), and medical history. Body weight and height were recorded to the nearest 0.1 kg and 0.1 cm, respectively, using calibrated scales. Body mass index (BMI) was calculated as weight [kg] divided by height [m^2^]. An inelastic tape measure was used to measure the waist circumference of the subjects while they were standing comfortably with equal weight on both feet. Waist circumference was measured in the horizontal plane midway between the iliac crest and the lower edge of the ribs. In participants without pacemakers, fat-free mass [kg] and fat mass [kg] were measured by bioelectrical impedance analysis (BIA) using a Nutriguard-M multifrequency device (Data Input, Pöcking, Germany) and NUTRI4 software (Data Input). Electrodes were placed on the hands, wrists, ankles, and feet. The test frequency was measured at 5, 50, and 100 kHz according to the manufacturer’s instructions. [1] Diabetes was defined as known medical diagnosis of diabetes mellitus or use of antidiabetic medication (Anatomic Therapeutic Chemical Classification System; code A10), or diabetes was defined as HbA1c ≥6.5% or non-fasting blood glucose ≥11.1 mmol/l [2]. Participants were asked to lie down and have blood drawn from the cubital vein. HbA1c was determined by high-performance liquid chromatography with a coefficient of variation of 1.5% and spectrophotometric detection (Diamat Analyzer; Bio-Rad, Munich, Germany). Serum concentrations of high-sensitivity C-reactive protein (hs-CRP) were measured by nephelometry on the Dimension VISTA (Siemens Healthcare Diagnostics, Eschborn, Germany). [3–5] Systolic and diastolic blood pressure measurements (HEM-705CP, Omron Corporation, Tokyo, Japan) were also taken three times on the right arm of the seated patients, with a 3-minute rest period after each measurement. Hypertension (systolic blood pressure ≥140 mmHg and/or diastolic blood pressure ≥90 mmHg) was defined as the mean of the second and third readings. History and/or echocardiographic signals of previous myocardial infarction (MI) provided information on previous MI [6].

---

**Table A1.** Sensitivity analyses excluding 149 cases with self-reported physician-diagnosed myocardial infarction: Results from linear regression models for associations of periodontitis parameters with cardiac measures (normalized to height^2.7^) using pooled data.

|  | BOP |  | Mean PD |  | Percentage of sites with PD ≥ 4mm |  | Mean CAL |  | Number of missing teeth |  |
| --- | --- | --- | --- | --- | --- | --- | --- | --- | --- | --- |
| Outcome variable | β (95% CI) | p | β (95% CI) | p | β (95% CI) | p | β (95% CI) | p | β (95% CI) | p |
| LV end-diastolic volume; mL/m<sup>2.7</sup> | 0.58 (-0.23; 1.39) | 0.161 | 0.38 (0.07; 0.69) | 0.016 | 0.01 (-0.00; 0.02) | 0.204 | 0.20 (0.05; 0.35) | 0.008 | 0.05 (0.02; 0.08) | 0.001 |
| LV end-systolic volume; mL/m<sup>2.7</sup> | 0.32 (-0.11; 0.74) | 0.141 | 0.17 (0.01; 0.33) | 0.042 | 0.01 (0.00; 0.01) | 0.060 | 0.10 (0.02; 0.17) | 0.015 | 0.03 (0.01; 0.05) | <0.001 |
| LV wall thickness; mm/m<sup>2.7</sup> | 0.04 (-0.02; 0.09) | 0.194 | 0.02 (0.00; 0.04) | 0.044 | 0.00 (-0.00; 0.00) | 0.110 | 0.00 (-0.00; 0.01) | 0.331 | 0.00 (0.00; 0.00) | 0.031 |
| LV mass; g/m<sup>2.7</sup> | 1.81 (0.10; 3.71) | 0.063 | 1.18 (0.45; 1.92) | 0.002 | 0.03 (-0.00; 0.05) | 0.051 | 0.50 (0.15; 0.85) | 0.005 | 0.16 (0.10; 0.23) | <0.001 |
| LV stroke volume; mL/m<sup>2.7</sup> | 0.26 (-0.42; 0.95) | 0.453 | 0.21 (-0.05; 0.48) | 0.111 | 0.00 (-0.01; 0.01) | 0.733 | 0.10 (-0.02; 0.23) | 0.103 | 0.02 (-0.00; 0.05) | 0.066 |
| LV ejection fraction; % | -0.68 (-1.99; 0.62) | 0.305 | -0.25 (-0.75; 0.25) | 0.328 | -0.02 (-0.03; -0.00) | 0.093 | -0.18 (-0.41; 0.06) | 0.151 | -0.04 (-0.09; -0.01) | 0.098 |
| E to E' ratio | 0.34 (0.13; 0.55) | 0.002 | 0.05 (-0.03; 0.13) | 0.224 | 0.00 (-0.00; 0.00) | 0.319 | -0.01 (-0.05; 0.03) | 0.546 | 0.00 (-0.01; 0.01) | 0.716 |
| Left atrium diameter; cm/m<sup>2.7</sup> | 0.01 (-0.01; 0.03) | 0.483 | 0.00 (-0.01; 0.01) | 0.536 | 0.00 (-0.00; 0.00) | 0.627 | 0.00 (-0.00; 0.01) | 0.116 | 0.00 (-0.00; 0.00) | 0.841 |

Models were adjusted for age, sex, school education, smoking status, known or diagnosed diabetes, dental visits within the last 12 months, fat mass, fat-free mass, myocardial infarction and study. Bold indicates results with p < 0.05.
Abbreviations: β, beta regression coefficient; BOP, Bleeding on probing; CAL, clinical attachment loss; CI, confidence interval; E to E', early (E) mitral inflow peak diastolic velocity to early (E') mitral annular peak diastolic velocity; LV, left ventricular; PD, probing depth.

**Table A2.**
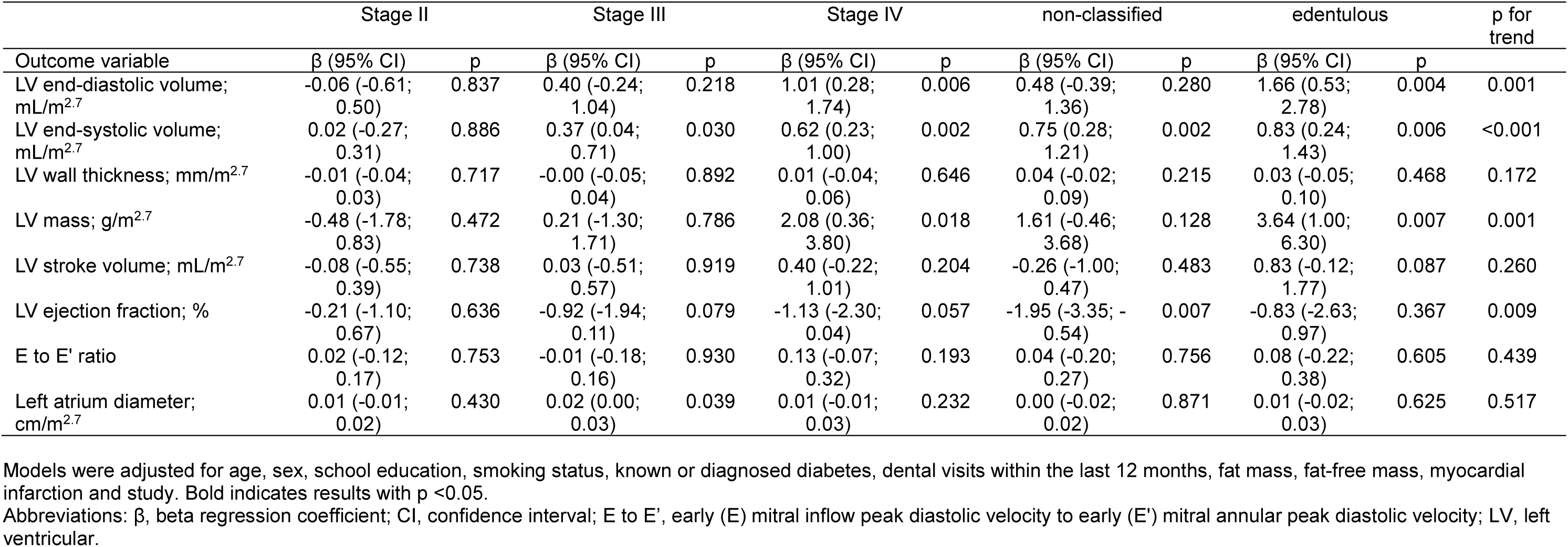
Sensitivity analyses excluding 149 cases with self-reported physician-diagnosed myocardial infarction: Results from linear regression models for associations of the 2018 classification (ref. “< stage II” (periodontal health, gingivitis or stage I periodontitis)) with cardiac measures (normalized to height^2.7^) using pooled data.

|  | Stage II |  | Stage III |  | Stage IV |  | non-classified |  | edentulous |  | p for trend |
| --- | --- | --- | --- | --- | --- | --- | --- | --- | --- | --- | --- |
| Outcome variable | $\beta$ (95% CI) | p | $\beta$ (95% CI) | p | $\beta$ (95% CI) | p | $\beta$ (95% CI) | p | $\beta$ (95% CI) | p | |
| LV end-diastolic volume; mL/m<sup>2.7</sup> | -0.06 (-0.61; 0.50) | 0.837 | 0.40 (-0.24; 1.04) | 0.218 | 1.01 (0.28; 1.74) | 0.006 | 0.48 (-0.39; 1.36) | 0.280 | 1.66 (0.53; 2.78) | 0.004 | 0.001 |
| LV end-systolic volume; mL/m<sup>2.7</sup> | 0.02 (-0.27; 0.31) | 0.886 | 0.37 (0.04; 0.71) | 0.030 | 0.62 (0.23; 1.00) | 0.002 | 0.75 (0.28; 1.21) | 0.002 | 0.83 (0.24; 1.43) | 0.006 | <0.001 |
| LV wall thickness; mm/m<sup>2.7</sup> | -0.01 (-0.04; 0.03) | 0.717 | -0.00 (-0.05; 0.04) | 0.892 | 0.01 (-0.04; 0.06) | 0.646 | 0.04 (-0.02; 0.09) | 0.215 | 0.03 (-0.05; 0.10) | 0.468 | 0.172 |
| LV mass; g/m<sup>2.7</sup> | -0.48 (-1.78; 0.83) | 0.472 | 0.21 (-1.30; 1.71) | 0.786 | 2.08 (0.36; 3.80) | 0.018 | 1.61 (-0.46; 3.68) | 0.128 | 3.64 (1.00; 6.30) | 0.007 | 0.001 |
| LV stroke volume; mL/m<sup>2.7</sup> | -0.08 (-0.55; 0.39) | 0.738 | 0.03 (-0.51; 0.57) | 0.919 | 0.40 (-0.22; 1.01) | 0.204 | -0.26 (-1.00; 0.47) | 0.483 | 0.83 (-0.12; 1.77) | 0.087 | 0.260 |
| LV ejection fraction; % | -0.21 (-1.10; 0.67) | 0.636 | -0.92 (-1.94; 0.11) | 0.079 | -1.13 (-2.30; 0.04) | 0.057 | -1.95 (-3.35; -0.54) | 0.007 | -0.83 (-2.63; 0.97) | 0.367 | 0.009 |
| E to E' ratio | 0.02 (-0.12; 0.17) | 0.753 | -0.01 (-0.18; 0.16) | 0.930 | 0.13 (-0.07; 0.32) | 0.193 | 0.04 (-0.20; 0.27) | 0.756 | 0.08 (-0.22; 0.38) | 0.605 | 0.439 |
| Left atrium diameter; cm/m<sup>2.7</sup> | 0.01 (-0.01; 0.02) | 0.430 | 0.02 (0.00; 0.03) | 0.039 | 0.01 (-0.01; 0.03) | 0.232 | 0.00 (-0.02; 0.02) | 0.871 | 0.01 (-0.02; 0.03) | 0.625 | 0.517 |

Models were adjusted for age, sex, school education, smoking status, known or diagnosed diabetes, dental visits within the last 12 months, fat mass, fat-free mass, myocardial infarction and study. Bold indicates results with p < 0.05.
Abbreviations: $\beta$ , beta regression coefficient; CI, confidence interval; E to E', early (E) mitral inflow peak diastolic velocity to early (E') mitral annular peak diastolic velocity; LV, left ventricular.

**Fig. A1.**
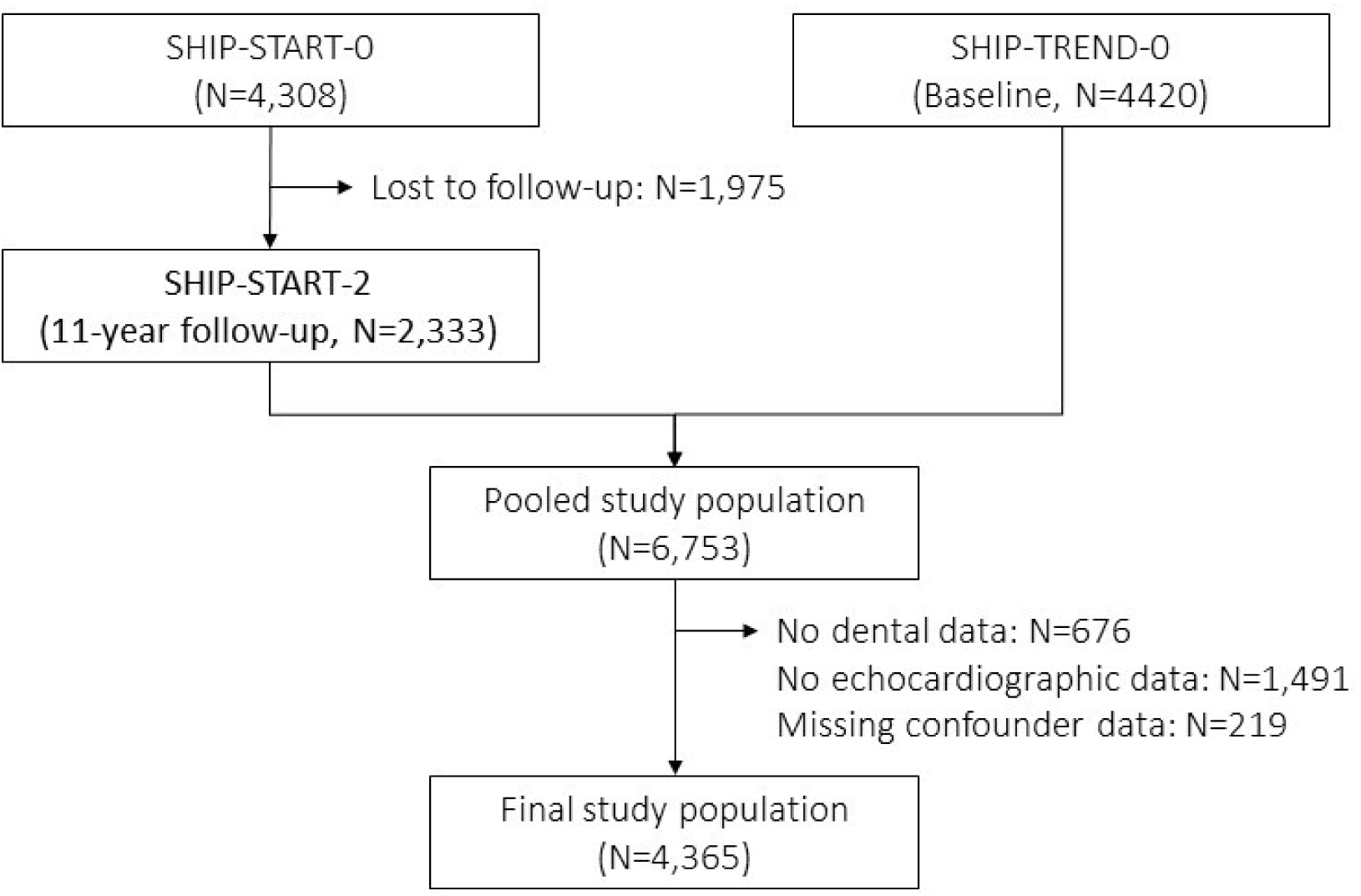
Flowchart of included and excluded participants from SHIP-START-2 and SHIP-TREND-0.

